# Noninvasive MRD monitoring and profiling of clonal evolution by ctDNA in patients with advanced cancers treated within molecular tumor boards

**DOI:** 10.64898/2026.05.27.26353937

**Authors:** Lavanya Ranganathan, Julia C. Kuehn, Christian Klingler, Thomas Pauli, Patrick Metzger, Sabine Bleul, Ulrike Philipp, Fabian Hummel, Samuel Weinschenk, Max Deuter, Julian Rapp, Christof Winter, Holger Sültmann, Ingeborg Tinhofer, Florent Mouliere, Justyna Rawluk, Nikolas von Bubnoff, Eva Dazert, Anna L. Illert, Alexandra Nieters, Julius Wehrle, Christoph Peters, Tilman Brummer, Anne Schultheis, Silke Lassmann, Cornelius Miething, Heiko Becker, Martin Werner, Melanie Boerries, Justus Duyster, MTB-FR Network, DKTK EXLIQUID consortium, Florian Scherer

## Abstract

Circulating tumor DNA (ctDNA) from blood plasma has emerged as a promising biomarker for noninvasive profiling of tumor mutational landscapes and disease monitoring across cancers. In this study, we developed a targeted next-generation sequencing approach to explore the role of ctDNA for comprehensive tumor genotyping, early response prediction, and characterization of clonal heterogeneity in patients with advanced and rare cancers treated within molecular tumor boards. We applied our technology to 157 plasma specimens from 57 patients at distinct disease milestones and detected tumor variants in 96% of baseline samples, with 65% of them harboring actionable aberrations. Longitudinal monitoring of baseline mutations in on-treatment plasma revealed that ctDNA dynamics were significantly associated with clinical outcomes and enabled early prediction of disease progression. Finally, we observed substantial clonal heterogeneity over time, identifying emerging mutations in all analyzed plasma samples obtained at progression, including potentially targetable variants for subsequent personalized therapies.

## Introduction

Comprehensive genomic profiling of tumor tissue by next-generation sequencing (NGS) technologies and the implementation of molecularly stratified therapies are transforming clinical management in oncology (1–3). While biomarker-informed targeted therapies have been successfully integrated into routine clinical practice for selected tumor entities such as non-small cell lung cancer (NSCLC) and melanoma, treatment options for the majority of cancer types still largely rely on conventional (immuno-)chemotherapy, particularly in the relapsed/refractory setting and in patients with rare tumors (4–6). Molecular tumor boards (MTBs) are widely established interdisciplinary forums that enable extended genomic profiling of tumors from patients with advanced and rare cancers after exhaustion of conventional treatment options, with the aim of identifying and initiating innovative targeted therapeutic strategies (7–13). Molecular testing in the MTB setting critically depends on the availability of adequate tumor tissue. However, broad and in-depth genomic characterization is frequently hampered by poor quality of archival biopsy material, insufficient tumor content, or limited accessibility of tumor lesions due to their anatomical location or patient’s clinical condition (14,15). In addition, longitudinal molecular profiling from tumor tissue remains challenging, as repeated biopsies at the time of disease progression are rarely obtained due to their invasive nature. Separately, monitoring of treatment response after initiation of MTB-recommended therapies is typically performed by radiologic imaging, which is often limited by suboptimal sensitivity and specificity for early response assessment and detection of measurable residual disease (MRD) (16).

Circulating tumor DNA (ctDNA) from blood plasma has emerged as a promising biomarker for noninvasive profiling of tumor mutational landscapes and longitudinal disease monitoring across oncology (17–20). In this study, we applied a custom targeted capture NGS approach to investigate the value of ctDNA for comprehensive tumor genotyping, early response prediction, and characterization of temporal clonal heterogeneity in patients with advanced cancers undergoing treatment within MTBs.

## Materials and Methods

### Patient cohort and study design

Patients included in this study were enrolled between October 2021 and October 2024 in the MTB of the University Medical Center Freiburg (MTB-FR) by their treating physician due to advanced disease with no remaining standard treatment options, the presence of a rare cancer entity, and/or young age at cancer diagnosis (*n*=51) (DRKS00025847). Molecular analyses and interdisciplinary treatment recommendations within the MTB-FR followed standard operation procedures as previously described (11,15). In brief, genomic profiling of tumor tissue was performed by applying diverse NGS-based technologies (i.e., panel sequencing, whole exome sequencing [WES]), followed by the identification of genetic alterations and complex molecular biomarkers by standardized bioinformatics workflows, as described before (11,15,21). Then, based on these results, personalized treatment recommendations were provided after interdisciplinary evaluation (11,15). Another subset of patients (*n*=6) included in this study was treated at other sites within MTBs after genetic characterization and interdisciplinary treatment recommendation. In this research study, we specifically included patients who harbored actionable genetic alterations identified by tumor sequencing and received personalized biomarker-guided treatment recommendations by the respective MTBs (**Suppl. Tables 1,2**). All included patients underwent either MTB-recommended targeted therapies or conventional bridging therapies (**Suppl. Table 1**). All patients provided written informed consent approved by the local ethics committee in accordance with the Declaration of Helsinki for both the enrollment in MTBs, and/or associated tumor analyses as well as for the participation in this research study, the collection of biospecimens and clinical data, and experimental analyses (DRKS00025847, ethic vote no. 369/19; DRKS00015849).

### Sample collection, processing, and sequencing

157 blood samples from all included patients (*n*=57) were collected at prespecified time points at MTB presentation (‘baseline’), during MTB-recommended therapy or bridging therapy (‘on-treatment’), and at disease progression. Cancer Personalized Profiling by Deep Sequencing (CAPP-Seq) was performed as previously described, applying a custom pan-cancer hybrid capture panel that covered 266 genes and 540 kb genomic space (**Suppl. Table 3**, Suppl. Methods). For tumor genotyping from ctDNA, somatic alterations were identified by paired analysis of baseline plasma with germline DNA, as described before (22–24). To monitor mutations detected from baseline ctDNA in on-treatment and other longitudinal plasma samples (MRD monitoring), we applied a previously described Monte Carlo framework to define ctDNA positivity/negativity (22–24). Specificity was assessed by monitoring of mutations derived from baseline ctDNA of patient’s plasma in cell-free DNA (cfDNA) from plasma samples of healthy individuals (Suppl. Methods) (22–24). Finally, ctDNA levels were quantified as allelic frequencies (AF) in percent. Further details are provided in the Suppl. Methods.

### Response criteria and outcome analyses

Clinical, pathological, radiographic, and laboratory data were assessed as part of routine clinical care. Response to MTB-recommended targeted or conventional bridging therapies was defined radiographically according to the RECIST criteria (25). Progression-free survival (PFS) was defined as the time from the start of therapy until disease progression, death from any cause, or last follow-up. For ctDNA-based response assessment during treatment, patients with at least one blood sample before the start of therapy and at least one sample during treatment (between 7 and 150 days after start of therapy) were included. We further specifically explored ctDNA dynamics from baseline to first on-treatment plasma samples and defined two groups: those with increasing and decreasing ctDNA concentrations. For these two groups, the best radiographic response as well as the radiographic response closest to the first on-treatment plasma samples were assessed.

### Statistical analysis

We used non-parametric Mann-Whitney *U* test to compare continuous variables and chi-square test for categorical variables. Linear relationships were determined using Pearson correlation coefficient. Time-to-event variables were visualized using the Kaplan-Meier method. Log-rank test was used to evaluate survival differences. Lead time was defined as the time from increasing ctDNA levels to radiographic disease progression. Statistical tests were performed using GraphPad Prism (version 9.1.1) and MedCalc (Version 20.111). *P*-values < 0.05 were considered as significant.

## Results

### Study cohort and patient characteristics

Fifty-seven patients with advanced and rare solid tumors with known actionable genetic alterations were included in this study. Fifty-one patients were identified within the MTB-FR, and an additional six patients were included from external MTBs, with a median age of 58 years (range: 23-84) (**Suppl. Fig. 1A, Suppl. Tables 1,2**). The patient cohort comprised ten distinct tumor entities, with lung (23%, *n*=13) and biliary tract (21%, *n*=12) cancers representing the most frequent tumor types (**Fig. 1A, Suppl. Tables 1,2**). Tumor tissue genotyping within MTBs using panel-based approaches or WES revealed 57 actionable aberrations, including *KRAS* G12C mutations, inactivating *BRAF* mutations, *IDH1/2* variants, and *BRAF* V600E mutations as the most common druggable alterations identified in this cohort, leading to case-specific MTB treatment recommendations (**Fig. 1A, Suppl. Table 1, Suppl. Figs. 2,3**). Following interdisciplinary assessment at the MTBs, a total of 66 therapies were initiated across all 57 patients, comprising targeted treatment in 53% and conventional bridging therapies such as immunochemotherapy or radiation in 47% of cases **(Fig. 1A, Suppl. Table 1)**.

**Figure 1.**
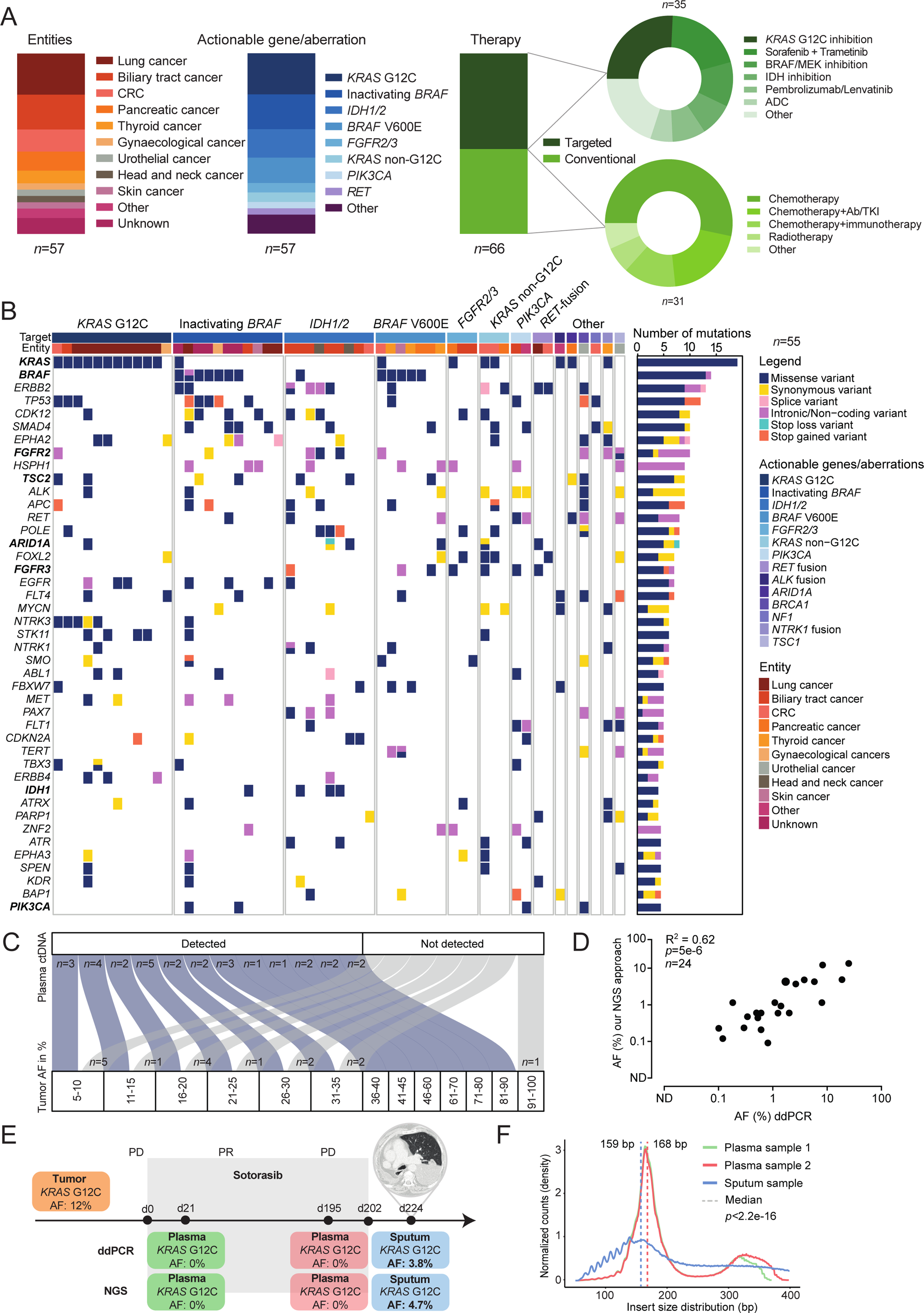
Cohort characteristics and noninvasive genotyping results. **(A)** Distribution of tumor entities (left), identified actionable mutations by tumor sequencing (middle), and therapies received after MTB recommendation (right) in the entire study cohort. CRC, colorectal cancer; ADC, antibody-drug conjugate; AB, antibody; TKI, tyrosine kinase inhibitor **(B)** Case-level mutational profiles based on noninvasive ctDNA based genotyping. Each column represents one case, each row represents a gene. Genes with at least 4 recurrent mutations in the cohort are shown. The number of alterations in each gene in this cohort is shown as a bar graph on the right. Tumor entity and actionable mutations/genes are depicted as a color code in the first two rows. Bold gene names indicate genetic alterations resulting in treatment recommendations. **(C)** Sankey plot visualizing the overall concordance of identified actionable SNVs between tumor and plasma-ctDNA genotyping (*n*=45). Of note, allele frequencies (AF) of actionable mutations found in 6 patients treated at other sites were not known. **(D)** Scatter plot showing the correlation of AFs measured by next-generation sequencing (NGS) and digital droplet PCR (ddPCR). **(E)** Timeline depicting the clinical course and biospecimens available from a patient with mucinous NSCLC with *KRAS* G12C mutation receiving sotorasib treatment. AFs for each sample type and method are shown. d, day; PD, progressive disease; PR, partial remission. **(F)** Fragment size distributions of two plasma samples and one sputum sample of the NSCLC case depicted in (E). Median size of each sample type is depicted as dashed line. bp, base pairs.

### Noninvasive tumor genotyping from ctDNA

Baseline plasma samples at initial MTB presentation were available from all 57 patients to explore the performance of noninvasive tumor genotyping from ctDNA by our targeted capture NGS approach (**Suppl. Fig. 1**). In 96.4% of cases (55/57), at least one genetic aberration was detected in baseline blood plasma, with a median of 7 mutations per sample and a range of 1-41 mutations (**Fig. 1B, Suppl. Fig 4A,B**). Most frequently mutated genes included *KRAS* (35%), *BRAF* (24%), *ERBB2* (23%), and *TP53* (22%), largely reflecting tumor tissue genotyping (**Fig. 1B**). We identified 65% of all actionable single nucleotide variants (SNVs) by plasma genotyping with a median AF of 0.5% (range: 0.05%-58%), whereas fusions were not detected in any of the cases noninvasively (**Fig. 1B, Suppl. Table 4**). Notably, SNVs with high allelic representation in tumor tissue (≥ 35%) could be detected in 92% of corresponding plasma samples, while tumor mutations ranging between 5% and 35% were found in 55% of baseline plasma samples, indicating a strong correlation of ctDNA detection rates with tumor burden **(Fig. 1C, Suppl. Fig 5**). To validate these findings, we performed digital droplet PCR (ddPCR) in a subset of baseline plasma specimens that are known to harbor *KRAS* G12C and *BRAF* V600E mutations (*n*=24), demonstrating a significant correlation between the AFs detected by ddPCR and our NGS-based technology (*p*=5e-6, R^2^= 0.62, **Fig. 1D**).

In one patient with mucinous adenocarcinoma of the lung, we never identified a known tumor-specific *KRAS* G12C mutation in blood plasma despite high tumor volumes and stage IV disease (**Fig. 1E**). However, this driver mutation was readily detectable in cfDNA isolated from a sputum sample obtained at disease progression, highlighting that the detectability of tumor-derived variants across biofluids can vary substantially depending on tumor entity, anatomical context, and tumor-specific biological features. Of note, we observed significant differences in cfDNA fragment size distributions across specimen types within this patient, with sputum showing a higher abundance of smaller fragments (median: 159 bp) compared to blood plasma (median: 168 bp) (**Fig. 1F**, *p*=2.2e-16).

### MRD monitoring and early response prediction by ctDNA profiling

In a next step, we investigated whether ctDNA monitoring during treatment following MTB recommendations could serve as a clinically useful MRD biomarker. A total of 59 on-treatment plasma samples were available from 22 patients for MRD monitoring **(Suppl. Fig. 1A**). The specificity of our monitoring approach was 96%, as determined by tracking patient-specific tumor variants from baseline specimens (*n*=55) in plasma samples obtained from 12 healthy individuals (Suppl. Methods) **(Fig. 2A, Suppl. Fig.6**).

**Figure 2.**
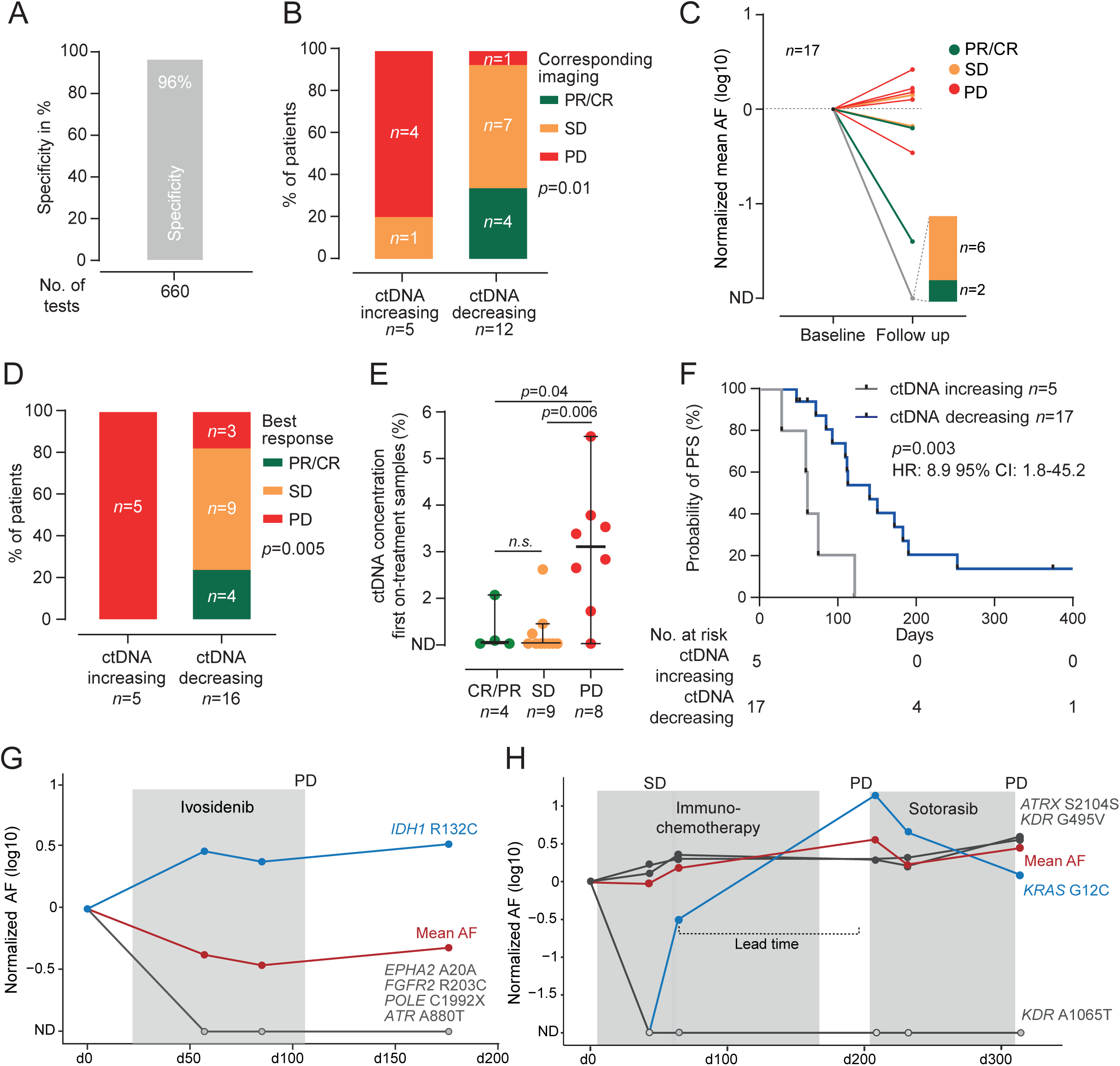
Monitoring of plasma ctDNA and early prediction of clinical outcomes. **(A)** Bar graph showing the specificity of the ctDNA monitoring approach. No., number. **(B)** Bar graphs depicting radiologic response in CT scans corresponding to the first on-treatment plasma sample obtained after initiation of treatment following MTB recommendation in those cases with increasing ctDNA levels compared to the baseline (left) or decreasing ctDNA levels (right). CR, complete remission; PR, partial remission; SD, stable disease; PD, progressive disease. **(C)** Dynamic changes of mean ctDNA levels in the first on-treatment plasma sample, normalized to the baseline plasma sample. Each line is colored according to the patient’s response in corresponding CT scans. ND, not detected; CR, complete remission; PR, partial remission; SD, stable disease; PD, progressive disease. **(D)** Bar graphs showing the best radiologic response in patients with increasing (left) or decreasing plasma ctDNA levels in the first on-treatment plasma samples. CR, complete remission; PR, partial remission; SD, stable disease; PD, progressive disease. **(E)** Comparison of ctDNA levels in the first on-treatment samples between patients with CR/PR (green), SD (yellow), or PD (red) as best radiologic responses. Bold lines represent the median, error bars show the 95% confidence intervals. n.s., not significant. **(F)** Kaplan-Meier estimates showing progression-free survival in patients with increasing (grey) and decreasing (blue) ctDNA levels on the first on-treatment plasma sample. **(G,H)** Representative cases demonstrating longitudinal ctDNA monitoring, normalized to the baseline time point. Blue lines show the actionable mutation, red lines highlight the ctDNA mean AFs, and grey lines demonstrate lost clones during treatment.

In a first step, we identified 17 patients with available on-treatment plasma samples obtained within a 40-day time period in relation to the first radiological assessment after initiation of treatment, to explore early ctDNA dynamics and correlation with radiological response. Five patients revealed increasing ctDNA levels and 12 showed decreasing ctDNA concentrations in their first on-treatment plasma samples (**Fig. 2B,C**). While 80% of patients with increasing levels showed progressive disease, 92% of patients with declining ctDNA concentrations had disease control with either partial response or stable disease on corresponding imaging (**Fig. 2B,C**).

Having shown associations between ctDNA and radiological response in contemporaneous scans, we then investigated whether early ctDNA dynamics could predict long-term and durable response as well as clinical outcomes following therapies initiated after MTB recommendations. A total of 21 patients with available on-treatment plasma samples were evaluable for the assessment of best radiological response over time. In this analysis, 81% of patients with declining ctDNA levels early into treatment showed durable disease control, whereas all patients with ctDNA concentrations exceeding the baseline ultimately developed disease progression (**Fig. 2D**). Further, concentrations of ctDNA in first on-treatment samples in patients with future radiological progression were significantly higher than those in patients with disease control (**Fig. 2E**). This was reflected in a significantly superior progression-free survival rate in patients with an early decrease of ctDNA levels (*p*=0.003, HR: 8.9, 95% CI: 1.8-45.2, **Fig. 2F**). Collectively, these findings suggest that serial ctDNA monitoring early during therapy could provide key advantages for response assessment in individual patients treated in the MTB setting. However, a subset of patients was still misclassified by ctDNA risk stratification, as they exhibited radiological disease progression despite declining mean ctDNA levels. One possible explanation for this phenomenon might be clonal heterogeneity under the selective pressure of targeted therapies, illustrated by a representative case highlighted in **Fig. 2G**. This patient showed decreasing ctDNA levels in most of the subclones early into treatment with ivosidenib, while the major *IDH1* R132C-carrying clone appeared to be refractory to this targeted treatment, leading to progressive disease in subsequent CT scans (**Fig. 2G**).

Next, we sought to explore the value of MRD monitoring for the prediction of cancer progression in individual cases. A total of 19 patients with available longitudinal plasma samples showed disease progression during the follow-up period. In 8 of these cases (42%), we found increasing ctDNA levels preceding radiological progression, with a median lead time of 53 days (range: 17-85 days) (**Suppl. Fig. 7**). The potential clinical utility of the monitoring approach could be illustrated in a patient with NSCLC, harboring a *KRAS* G12C mutation who received immunochemotherapy as bridging to sotorasib (**Fig. 2H**). A slight decrease of the mean AF in the first on-treatment plasma sample was reflected by stable disease in a corresponding CT scan. However, ctDNA levels in the following plasma specimen were increasing, predicting radiographic disease progression with a lead time of 77 days (**Fig. 2H**).

### Characterization of temporal clonal heterogeneity by ctDNA profiling

Among the most promising clinical applications of ctDNA is its potential use for the detection of clonal heterogeneity and resistance mechanisms in response to treatment, especially in situations where repetitive biopsies are challenging. To investigate clonal emergence noninvasively over time, we identified 16 patients with paired baseline and progression plasma samples (**Suppl. Fig. 1A**). We observed substantial temporal clonal evolution, with in median 77% unique mutations present either in the baseline (i.e., ‘lost variants’, median of 32%) or progression samples (i.e., ‘emerging variants’, median of 42%) alone, while the fraction of shared mutations (i.e., ‘truncal variants’) was in median 24% (**Fig. 3A**). Notably, we observed a higher number of unique aberrations in patients receiving conventional bridging therapies after MTB recommendation compared to those undergoing targeted treatment (**Fig. 3B**). However, when normalizing the number of unique mutations to therapy duration, no significant differences were observed between the two treatment modalities (**Fig. 3B**). Notably, the distribution of truncal and unique variants varied substantially across genes and within individual patients, with aberrations in *KRAS, TP53,* and *BRAF* being commonly preserved longitudinally and towards disease progression (**Fig. 3C,D, Suppl. Fig. 8**).

**Figure 3.**
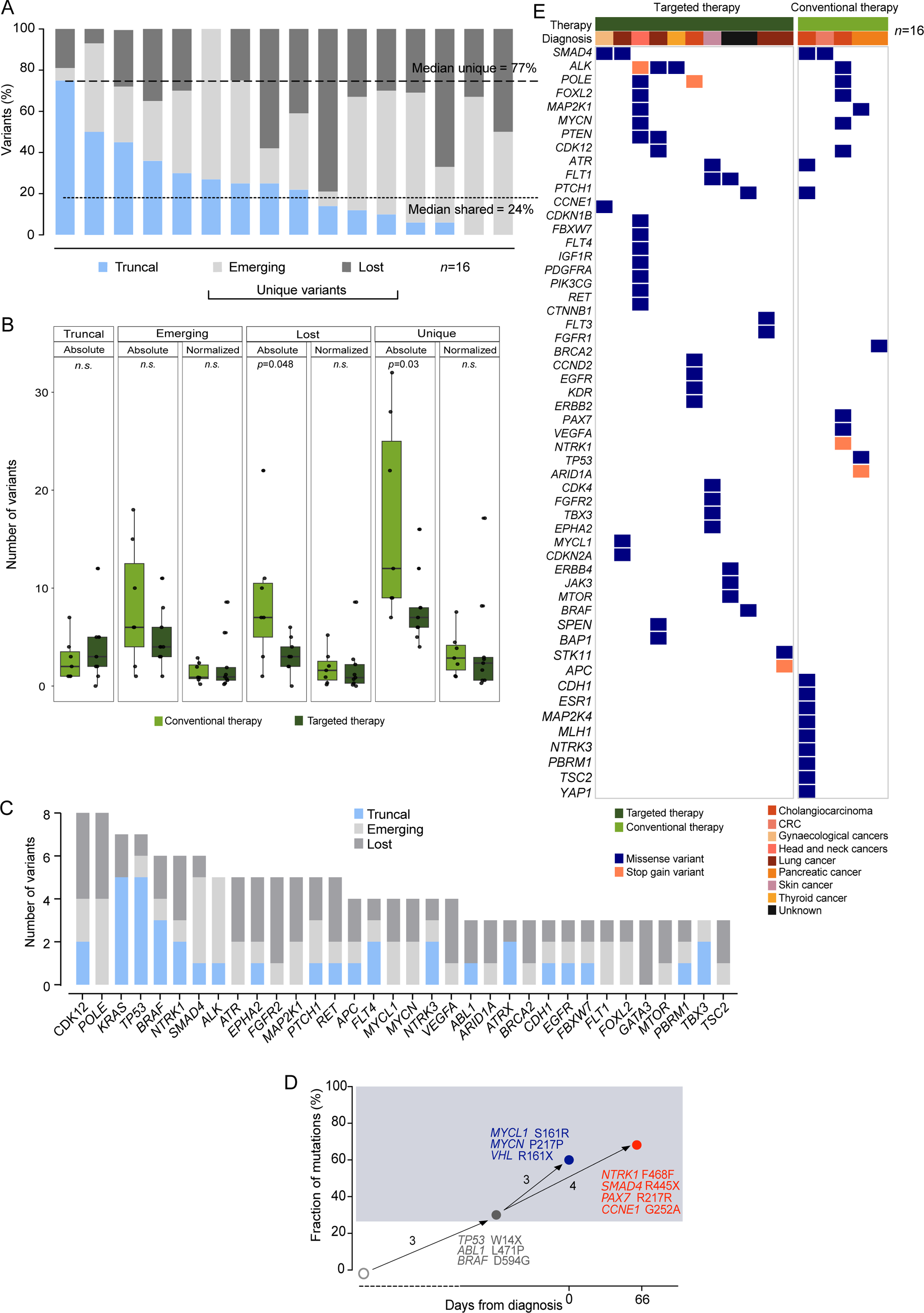
Clonal heterogeneity identified by ctDNA profiling. **(A)** Bar graphs depicting the proportion of truncal (blue), emerging (light grey), and lost variants (dark grey) for each patient with paired plasma at baseline and disease progression. **(B)** Number of truncal, emerging, lost, and unique (emerging plus lost) genetic aberrations detected in paired plasma samples obtained at baseline and progression for patients receiving targeted therapies (dark green) or conventional bridging therapies (light green) following MTB recommendations, both the absolute numbers (left) and normalized to treatment duration (right). Bold lines represent the median, error bars show the 95% confidence intervals. n.s., not significant **(C)** Bar graphs showing the cumulative number of truncal (blue), emerging (light grey), and lost variants (dark grey) per gene at the progression time point across the entire cohort of 16 patients with paired baseline-progression samples. **(D)** One representative case highlighting clonal evolution over time. The initial branch represents truncal mutations (grey) observed in both plasma samples at baseline and progression. At disease progression, three mutations depicted in blue were lost, while four mutations emerged (red) 66 days after the start of therapy. The x-axis represents treatment duration, with ‘0’ representing the baseline time point. The y-axis represents the fraction of mutations. **(E)** Case-level profiles of emerging variants at disease progression. Each column represents one case, each row represents a gene. Tumor entity and type of therapy are depicted as a color code in the first two rows.

Emerging mutations over time are of particular interest, as they could reflect resistance mechanisms to initiated therapies and represent actionable targets for subsequent treatment. Across all 16 patients, we observed at least one emerging genetic aberration at disease progression by ctDNA profiling, with *SMAD4* and *ALK* genes being most frequently affected in 25% of cases (**Fig. 3E**). To evaluate a potential clinical relevance of those newly identified variants and to investigate whether these alterations could guide further biomarker-driven targeted therapeutic options, we first assessed their pathogenicity based on the evaluation criteria of the MTB-FR (Suppl. Methods). Applying the REVEL score, 19% of all emerging variants were identified as ‘pathogenic’ (14/72, REVEL score 0.7-1.0), while 81% were classified as either ‘benign’ or ‘uncertain’ (**Fig. 4A, Suppl. Table 5**). The proportion of pathogenic aberrations did not differ between treatment modalities, irrespective of whether they emerged in response to targeted or conventional therapies (**Suppl. Fig. 9**). Further, four emerging mutations occurred in protein kinase domains and a total of four aberrations (29% of pathogenic variants) were classified as potentially treatment-guiding in the respective tumor entity (**Fig. 4A,B, Suppl. Table 5**). Overall, 25% (4/16) of patients with cancer progression following therapies implemented after MTB assessment could potentially benefit from noninvasive ctDNA genotyping to guide subsequent biomarker-driven treatment.

**Figure 4.**
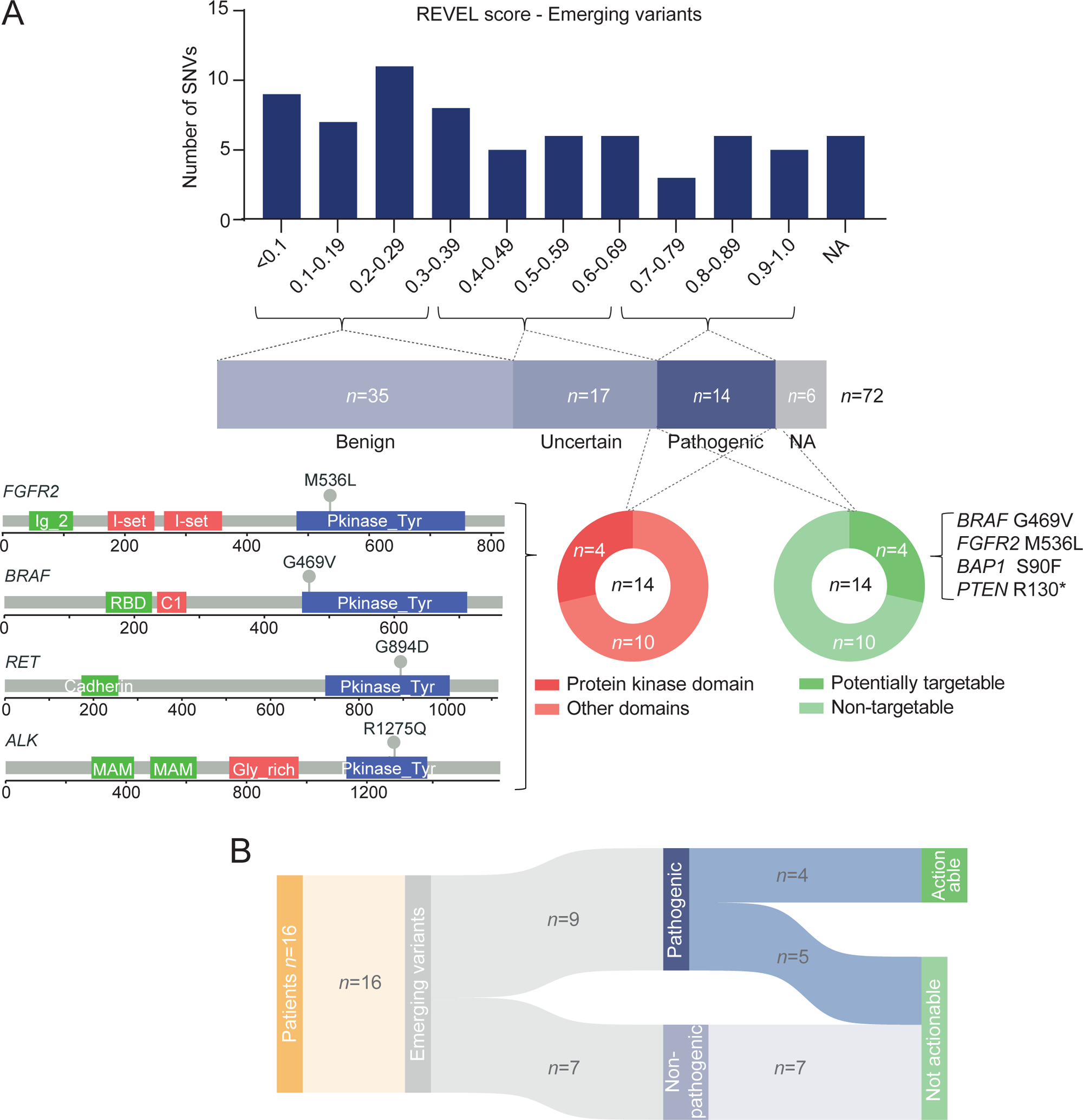
Characteristics of emerging ctDNA variants. **(A)** Distribution of the REVEL score of emerging variants (upper panel) and resulting classification in benign, uncertain, and pathogenic (middle panel). The lower panel highlights the genomic localization of pathogenic variants affecting the protein kinase domain (lower panel). Further, potentially targetable pathogenic aberrations are shown in dark green. **(B)** Sankey plot visualizing the classification of emerging variants at disease progression.

Together, our data highlight the potential of longitudinal ctDNA profiling to robustly capture clonal evolution and emerging actionable alterations that may inform biomarker-driven treatment in situations where repeated tumor biopsies are typically not available.

## Discussion

We here describe the value of high-throughput molecular profiling of ctDNA for noninvasive genotyping, assessment of early treatment response, and characterization of temporal clonal heterogeneity in patients with advanced and rare cancers treated within MTBs. Previous studies have highlighted the importance of ctDNA-based identification of actionable genetic aberrations for personalized treatment guidance in solid neoplasia (9,26,27). Yet, its broader clinical utility across diverse scenarios in the off-label MTB setting remains incompletely defined. Here, we developed a targeted liquid biopsy NGS technology enabling both sensitive longitudinal disease monitoring and comprehensive genotyping of actionable aberrations at baseline and clonal evolution over time. Our findings highlight three key insights with potential to improve clinical management of cancer patients treated within MTBs:

First, at MTB presentation, our approach facilitated robust noninvasive detection of targetable genetic alterations in blood plasma, achieving a detection rate of 65% across diverse cancer types, consistent with prior studies (19,28). Importantly, the sensitivity of ctDNA-based genotyping was closely associated with the allelic representation of mutations in corresponding tumor tissue, showing detection rates of up to 92% in cases with tumor AFs of at least 35%. These findings underscore that tissue-based genotyping remains the preferred modality for identifying treatment-guiding genetic aberrations. However, clinical scenarios such as insufficient tissue availability or limited accessibility of tumor lesions require alternative approaches. In such contexts, molecular profiling from blood plasma by ctDNA provides a reliable and clinically meaningful strategy for noninvasive identification of actionable mutations in the MTB clinical setting.

The second major finding of our study was the strong prognostic value of ctDNA and the association of early ctDNA dynamics with clinical outcomes during targeted or conventional bridging MTB therapy. Our results revealed that ctDNA monitoring early into treatment was predictive of subsequent disease progression. Notably, all patients who exhibited an increase of ctDNA levels at the first on-treatment time point experienced future radiographic progression, with a median lead time of 53 days. These findings may have important implications for treatment stratification and patient management in the MTB setting. For example, early molecular treatment failure by ctDNA might motivate an increased frequency of radiographic surveillance to enable earlier detection of clinical progression and timely modification of therapeutic approaches. Moreover, early noninvasive identification of treatment failure could help to avoid the continued use of ineffective off-label therapies, facilitating the initiation of alternative treatment strategies at a stage when patients are more likely to benefit and obviating unnecessary potential morbidity and financial toxicity.

Finally, we demonstrated substantial temporal clonal evolution by noninvasive ctDNA profiling in patients with progressive disease, with novel mutations emerging at the time of disease progression in all analyzed cases. Importantly, a subset of these emerging alterations was classified as pathogenic and represented potential targets for subsequent personalized therapies. These findings highlight the potential of our liquid biopsy approach to enable the real-time identification of resistance mechanisms and dynamic therapeutic vulnerabilities. This is particularly relevant in clinical settings like the MTB, in which repeated tumor biopsies are often unavailable or not feasible.

One particular strength of our study is the robust performance of both ctDNA genotyping and longitudinal monitoring in the real-world MTB setting, extending across diverse cancer entities rather than being restricted to a single tumor entity. However, our work harbors several limitations, and important challenges remain to be addressed. First, despite the overall breadth of the cohort, sample sizes were relatively small in some experimental subgroups, and the single-center design may limit generalizability of our findings. Second, none of the six fusions identified in tumor tissue were detected in plasma ctDNA, despite substantial efforts and the implementation of multiple fusion-calling algorithms. This reflects a well-recognized limitation of ctDNA-based technologies, as fusion events may be represented at very low levels in plasma and are generally more challenging to detect than SNVs. Finally, some of the actionable alterations and corresponding targeted therapies investigated in our study, such as *KRAS* G12C and the KRAS inhibitor sotorasib, have since entered routine clinical practice in defined indications, whereas they were still applied as off-label therapies at the time this study was conducted. This reflects the rapidly evolving therapeutic landscape of precision oncology and underscores the role of real-world MTB programs and corresponding translational research initiatives as early implementation platforms for molecularly-guided treatment strategies before their broader adoption into standard clinical care.

In summary, we developed a noninvasive ctDNA-based NGS technology for comprehensive profiling and longitudinal monitoring of genetic aberrations in patients with advanced cancers treated within an MTB framework. Our data support the utility of this strategy for the robust identification of actionable and personalized therapeutic targets at baseline and disease progression, the dynamic assessment of molecular treatment response, as well as early prediction of treatment failure and adverse clinical outcomes. Collectively, these findings highlight the potential of ctDNA profiling to complement tissue-based molecular diagnostics in selected situations and to support individualized treatment stratification in real-world precision oncology.

## Acknowledgements

The project was supported by the Berta-Ottenstein Programm Förderlinie Clinician Scientist (to J.C.K.), the Advanced Clinician Scientist Program of the Deutsche Gesellschaft für Innere Medizin (to F.S.), the Exzellenzstipendium of the Else Kröner-Fresenius-Stiftung (to F.S.), research grants of the Deutsche Forschungsgemeinschaft (SCHE 1870/3-1, to F.S.), the Mertelsmann Foundation (to F.S. and H.B.), the German Cancer Research Center (DKTK, to F.S. and H.B.), the Else Kröner-Fresenius-Stiftung (to F.S. and H.B.), and the Federal Ministry of Research, Technology and Space (BMFTR, to H.B.). The work on fragmentation profiling was supported and performed as a part of the Scientific Exchange Grant (SEG No.: 10693) from European Molecular Biology Organization (EMBO, to L.R). The work was further supported by the Deutsche Forschungsgemeinschaft – CRC1479 (Project ID 441891347-S1 to M.B., T.P.), CRC 1160 (Project ID 256073931-Z02 to M.B:), CRC1453 (Project ID 431984000-S1 to M.B.), TRR167 (Project ID 259373024-Z01 to M.B.), TRR 353 (Project ID 471011418-SP02 to M.B.), TRR417 (Project ID: 540805631-TP S03 to M.B.), FOR 5476 UcarE (Project ID 493802833-P7 to M.B.) and the German Federal Ministry of Research, Technology and Space (BMBFTR) within the National Decade against Cancer program for PM4Onco–FKZ 01ZZ2322A (M.B., P.M., T.P.) and for SATURN3 (01KD2206L to M.B.). We received also further funding by the Comprehensive Cancer Center Freiburg (CCCF), core support for molecular diagnostics in and by the Institute for Surgical Pathology and the local steering committee of DKTK partner site Freiburg. We further thank the team of the Genomics and Proteomics Core Facility, German Cancer Research Center/DKFZ, Heidelberg, Germany for their sequencing service, and the German Cancer Consortium (DKTK) and FREEZE/FREEZE-O Freiburg for biobanking. We further thank the ‘Centers for Personalized Medicine (ZPM)’ network Baden-Wuerttemberg, Germany.

## Author Contributions

Conception and design: F.S., C.W., H.S., I.T., N. vB.

Administrative support: S.B., U.P., A.N, C.P.

Provision of study materials or patients: J.C.K, A.L.I., S.L., A.S., M.W., C.M., H.B., J.D., M.B., F.S.

Collection and assembly of data: L.R., J.C.K, T.P., U.P., S.W., F.S.

Data analysis and interpretation: L.R., J.C.K, C.K., T.P., P.M., J.R., F.H., S.W., M.D., T.B., S.L., A.S., M. B., F.S.

Manuscript writing: All authors

Final approval of manuscript: All authors Accountable for all aspects of the work: All authors

## Data availability

Pseudonymized clinical and demographic data for cases considered in this study, as well as tumor mutational data and other relevant data are provided in the Supplementary Data. Owing to restrictions related to the dissemination of germline sequence information included in the informed consent forms used to enroll study subjects in the MTB-FR observational study, we are unable to provide access to raw sequencing data. Reasonable requests for additional data will be reviewed by the authors to determine whether they can be fulfilled in accordance with these privacy restrictions.

## Code availability

The analyses were conducted using existing software and tools, which are detailed within the Methods section. Should there be any inquiries regarding the computational methods employed, please contact the corresponding author for further information.

## Supplementary Information

This manuscript contains additional supplementary information in the Data Supplement.

