## Supplementary material for "Noninvasive MRD monitoring and profiling of clonal evolution by ctDNA in patients with advanced cancers treated within molecular tumor boards": Data Supplement

**Data Supplement for**

### Table of contents

**Supplemental Methods**

**Supplemental References**

**Supplemental Figures**

**Supplemental Tables – attached separately**

**Supplemental Table 1.** Case-level patient characteristics.

**Supplemental Table 2.** Clinical characteristics of the entire cohort.

**Supplemental Table 3.** NGS panel design (hg19).

**Supplemental Table 4.** SNVs detected in baseline plasma.

**Supplemental Table 5.** Emerging variants detected in blood plasma at progression.

### Supplemental Methods

Sample collection and processing

Blood samples of all included patients were centrifuged to obtain plasma and plasma-depleted whole blood (PDWB). Cell-free DNA (cfDNA) from the plasma compartment and cellular DNA from PDWB were extracted as previously described (1,2). Briefly, plasma samples were used for cfDNA isolation using the QIAmp circulating cell-free nucleic acid kit (Qiagen). The median plasma volume was 7.7 mL (range: 1.34 - 9.13 mL). PDWB was used for cellular DNA extraction using the QIAmp DNA Mini kit (Qiagen) as per the manufacturer's protocol. Cellular DNA was then sheared to a fragment size of 300 bps using the Covaris M220 instrument. DNA was quantified using either the Qubit dsDNA High Sensitivity kit (Thermo Fischer Scientific) for cfDNA Qubit or the dsDNA Broad Range kit (Thermo Fischer Scientific) for cellular DNA. Cellular DNA from PDWB was used as germline control.

One sputum sample was available from one patient with lung cancer. This sample was centrifuged at 1600 x g and supernatant was separated from the debris. Then, cfDNA extraction was performed using the QIAamp® DSP circulating NA Kit (Ref.No. 61504; QIAGEN, Hilden, Germany). The sample was distributed over 3 columns to prevent clogging and the resulting eluates were pooled. To qualitatively assess the presence of cellular DNA contamination in sputum-cfDNA, an electrophoresis-based quantification using the TapeStation 2000 (Agilent) assay was performed. The cell-free fraction of the sputum sample contained no high molecular weight DNA, the median fragment size in the TapeStation analysis was 159 bp.

In addition to the patient cohort, plasma samples were collected from healthy individuals (*n*=24). The healthy individual cohort was divided into two sub-cohorts. One cohort of 12 healthy plasma samples was used for background correction (see below), while the second cohort of healthy plasma samples (*n*=12) was used to test the specificity of our ctDNA based monitoring approach, as reported previously (see below for details) (3,4). Plasma samples from healthy controls were treated similarly to the patient plasma samples.

<u>Sequencing panel design and library preparation</u>

Genes previously reported to be relevant in solid cancers were included to design a custom targeted capture sequencing panel backbone, including genes of the MSK-IMPACT panel and a previously reported pan-solid cancer panel (5,6). The size of this panel backbone was 523 kb. To capture known resistance mutations, candidate regions were identified based on a comprehensive literature review of *in vivo* studies, *in vitro* studies, and clinical trials focusing on resistance mechanisms in patients receiving targeted therapies for solid cancers (7–22). Resistance mutation targets spanning 17 kb genomic space were then added to the panel backbone for a total of 540 kb, covering 266 genes (**Suppl. Table 3**). This panel was used for the CAPP-Seq (Cancer Personalized Profiling by Deep Sequencing) pipeline that included library preparation and hybridization, using the KAPA HyperPrep Kit (Roche Sequencing Solutions) and xGen Hybridization and Wash kit (IDT), of all plasma and sheared germline samples. Biotinylated oligonucleotides were designed and obtained from IDT (hg19, GRCh37). Median cfDNA mass used for library preparation was 32 ng, median mass of cellular DNA from germline samples was 100 ng.

Next-generation sequencing and bioinformatics analyses

After hybridization-based sequence enrichment, high-throughput sequencing was performed using 2x150 bp paired end sequencing on the Illumina NextSeq 1000/2000 platform. After collapsed read duplicated, we obtained median depths of 1364x for plasma and 1372x for germline samples. The CAPP-Seq bioinformatics workflow was applied to identify somatic mutations by paired analysis of baseline plasma and germline (i.e., 'genotyping'), and to monitor circulating tumor DNA (ctDNA in plasma samples (i.e., 'monitoring'), as described before (1). Somatic nucleotide polymorphisms (SNP) were filtered by matched germline analysis and stereotypic background was removed using cfDNA from the first set of 12 healthy individuals, as previously described (1,4). FACTERA was utilized for the identification of fusions (23).

Noninvasive tumor genotyping and ctDNA monitoring

For genotyping from baseline plasma, we considered mutations above a threshold of 0.05 % variant allele frequency (VAF). In addition, as clonal hematopoiesis of indeterminate potential (CHIP) mutations may add background noise to ctDNA-based genotyping and MRD monitoring, we set a filter to remove variants identified in canonical CHIP genes which include mutations in *DNMT3A*, *TET2*, *ASXL1*, *ASXL2*, *CHEK2*, *ATM*, *JAK2*, *GNAS*, *SF3B1*, *U2AF1*, *RUNX1*, *SH2B3*, *STAT3*, *ETV6*, *EZH2*, *FLT3*, *CREBBP*, *CEBPA*, *CSF1R*,

*EP300*, *FOXP1*, *KIT*, *MPL*, *MYD88*, *NF1*, *NOTCH1*, *NOTCH2*, *PTEN*, *PTPN11*, *SETBP1*, *SETD2*, *SRSF2*, and *STAG2* (24,25).

For MRD monitoring using a previously described Monte Carlo framework, variant lists obtained from genotyping of baseline plasma samples were used to monitor ctDNA levels in matched longitudinal plasma samples from the same patient (1,26,27). Specificity of the monitoring approach was assessed by tracking each individual variant list derived from patient baseline cfDNA genotyping in the second set of 12 plasma samples from healthy donors. This resulted in 684 tests to assess specificity. Specificity was then calculated as (**Suppl. Fig. 6**):

$$\left(1 - \frac{number\ of\ false\ positives\ identified\ while\ monitoring\ in\ healthy\ samples}{total\ number\ of\ tests}\right) \times 100$$

$$= \left(1 - \frac{25}{660}\right) \times 100 = 96.21\%$$

A patient plasma sample was defined as ctDNA positive or ctDNA negative based on the Monte Carlo *p*-value threshold that was set according to the *p*-value at 96% in the specificity analysis. Importantly, these 12 cfDNA samples from healthy individuals were withheld for the purpose of specificity assessment, and not part of any filtering step described above. Concentrations of ctDNA were quantified as mean allele frequencies (AF) in percent. To define ctDNA increase/decrease in on-treatment plasma samples, the mean AF of monitored samples was normalized to the mean AF of the baseline sample.

<u>Clonal heterogeneity analysis</u>

Matched plasma samples obtained at progression and baseline time points were explored for the characterization of clonal heterogeneity over time. Somatic variants in both specimens were identified by noninvasive genotyping as described above. Then, the integrated set of variants identified in these samples were used to create a reporter list that was monitored in all serial plasma samples obtained from the patient, applying the same monitoring approach as described above. For clonal heterogeneity analysis, mutations were categorized into 3 groups: truncal mutations, which were observed in both pretreatment and progression time points; lost mutations that were only present in pretreatment samples; and emerging mutations, which were exclusively observed in the plasma specimen obtained at progression.

<u>Fragment size measurements</u>

In one NSCLC patient, we compared the fragment size distribution of paired plasma and sputum samples. For this analysis, FASTQ files were obtained, and adapters were trimmed using *trimgalore*. Then, adapter-trimmed FASTQ files were mapped using *bwa mem*. PCR duplicate-removed BAM files were then used for calculating the fragment size distributions across each sample. Fragment sizes were calculated using *Rsamtools* and plotted using *ggplot2* in an R environment running under version 4.3.2.

<u>Digital droplet PCR</u>

To validate the allelic representation of ctDNA across different technologies, we applied digital droplet PCR (ddPCR) assays, targeting *KRAS* p.G12C (c.34G>T) and *BRAF* p.D594G (c.1781A>G), to a subset of cfDNA samples. Isolation of cfDNA was performed as described above. All samples were analyzed in quadruplicates. Each reaction well contained 11 $\mu$L of ddPCR Supermix for Probes (Bio-Rad), 1.1 $\mu$L each of mutant- and wild-type-specific LNA probes (final concentration 250 nM per probe), and 0.22 $\mu$L each of forward and reverse primers (final concentration 900 nM per primer). For the sputum sample, 2 $\mu$L of cfDNA template at a concentration of 40 ng/$\mu$L was added per well, whereas for plasma samples, 7 $\mu$L of cfDNA was used per well irrespective of the cfDNA concentration. Nuclease-free water was added to each reaction to a final volume of 22 $\mu$L. Droplets were generated using the Automated Droplet Generator (Bio-Rad) and subsequently subjected to PCR cycling under established conditions (28). Droplet fluorescence was measured using the QX200™ Droplet Reader (Bio-Rad). Data analysis was performed with QX Manager Software v2.2 (Bio-Rad), and mutant and wild-type copy numbers per milliliter of plasma, as well as the AF (%) were calculated.

| ***KRAS* G12C** | | |
|---|---|---|
| **Primer** | **Direction** | **Sequence** |
| G12C | Sense | 5'- GGA TCA TAT TCG TCC ACA A -3' |
| | Anti-Sense | 5'- CCT GCT GAA AAT GAC TGA A -3' |
| **Probes** | **Variant** | **Sequence** |
| G12C | wild type | 5'- /5HEX/TAC +GC+C A+CC +AGC TC/3IABkFQ/ -3' |
| | c.34 G>T | 5'- /56-FAM/TAC +GC+C A+CA +AGC TC/3IABkFQ/ -3' |

| ***BRAF* D594G** | | |
|---|---|---|
| **Primer** | **Direction** | **Sequence** |
| D594G | Sense | 5'- CACCTCAGATATATTTCTTCATG- 3' |
| | Anti-Sense | 5' CACTCCATCGAGATTTCA- 3' |
| **Probes** | **Variant** | **Sequence** |
| D594G | wild type | 5'- /5HEX/ AGA+CCA+AAA+TCA+CCT ATT T /3IABkFQ/ -3' |
| | c.1781A>G | 5'- /56-FAM/ AGA+CCA+AAA+CCA+CCT ATT T /3IABkFQ/ -3' |

Data availability

Anonymized clinical and demographic data on all cases considered in this study, as well as genotyping and other relevant data are provided in the Supplementary Tables. Owing to restrictions related to dissemination of germline sequence information included in the informed consent forms used to enroll study subjects, we are unable to provide access to raw sequencing data. Reasonable requests for additional data will be reviewed by the senior authors to determine whether they can be fulfilled in accordance with these privacy restrictions. Requests for additional materials related to this work should be directed to F.S.

### Supplemental Figures

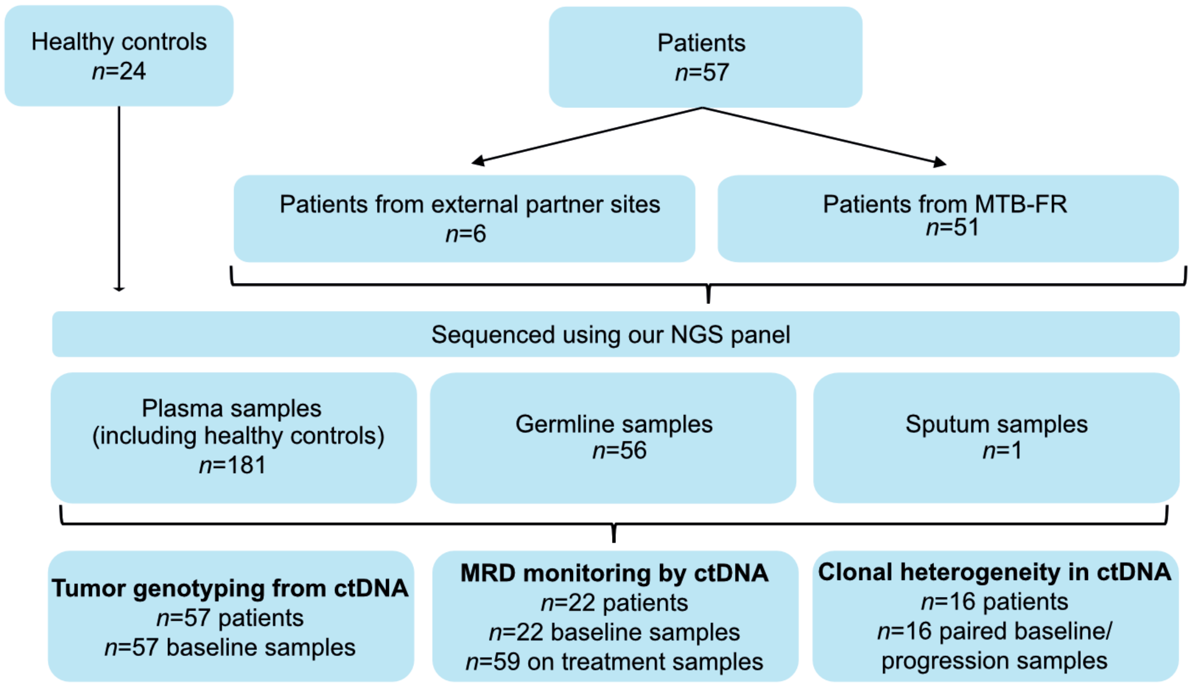


**Supplemental Figure 1. Overview of the study cohorts and sample usage.** MTB-FR, Molecular tumor board Freiburg; ctDNA, circulating tumor DNA; MRD, measurable residual disease.

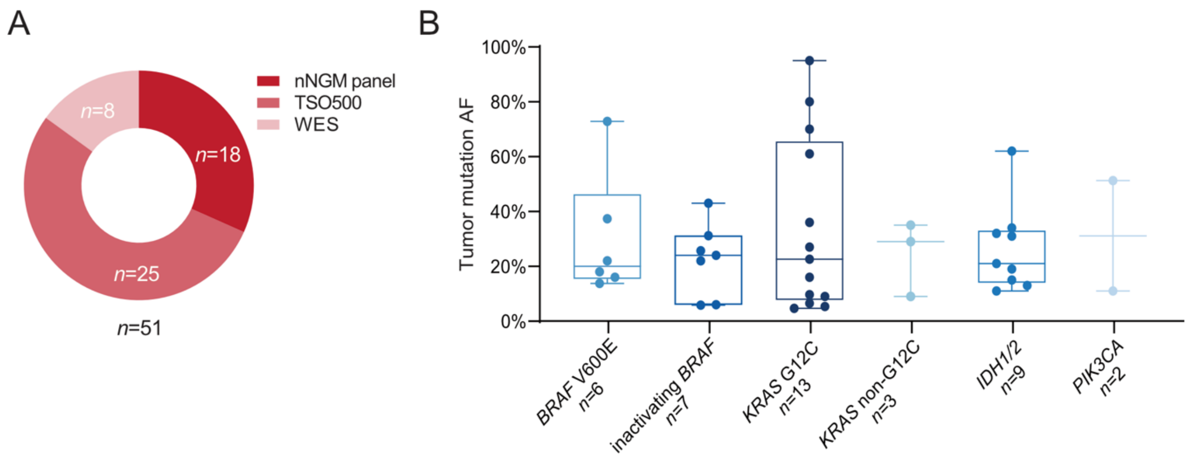


**Supplemental Figure 2. Genotyping of tumor tissue within MTBs. (A)** Circle plot showing the NGS methods used for tumor genotyping. nNGM, nationale Netzwerk Genomische Medizin; TSO500, TruSight Oncology 500; WES, Whole exome sequencing. **(B)** Distribution of allele frequencies (AF) of actionable tumor mutations identified in tumor tissue. Detection threshold of tumor genotyping was 5%. Boxes show the median and the 95% confidence intervals. Error bars represent range.

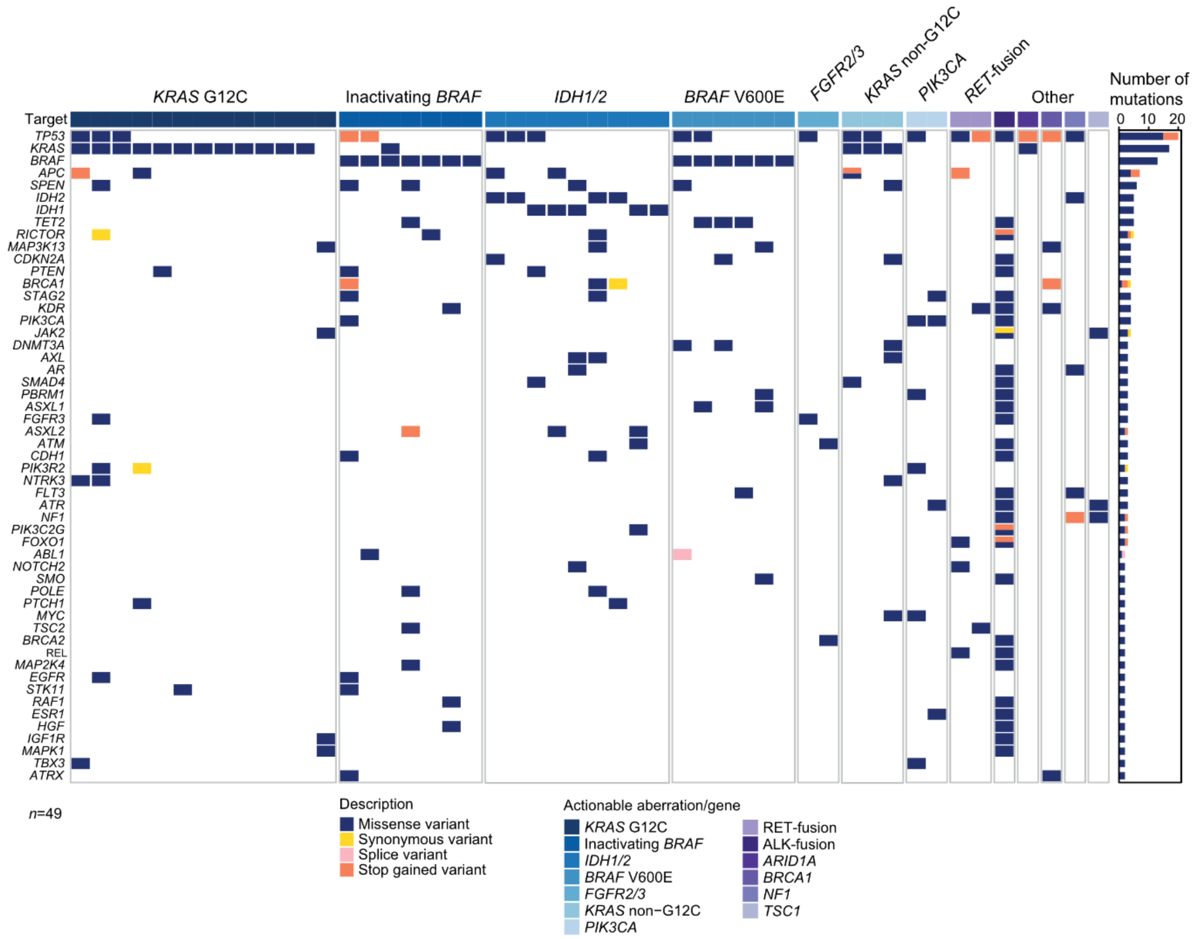


**Supplemental Figure 3. Tumor mutational profiles.** Case-level tumor mutational profiles based on NGS genotyping of tumor samples within MTBs. Each column represents one case, each row represents a gene. Genes with at least two recurrent mutations are shown. The number of alterations in each gene in this cohort is shown as a bar graph on the right. Tumor entity and actionable mutations are depicted as a color code in the first row.

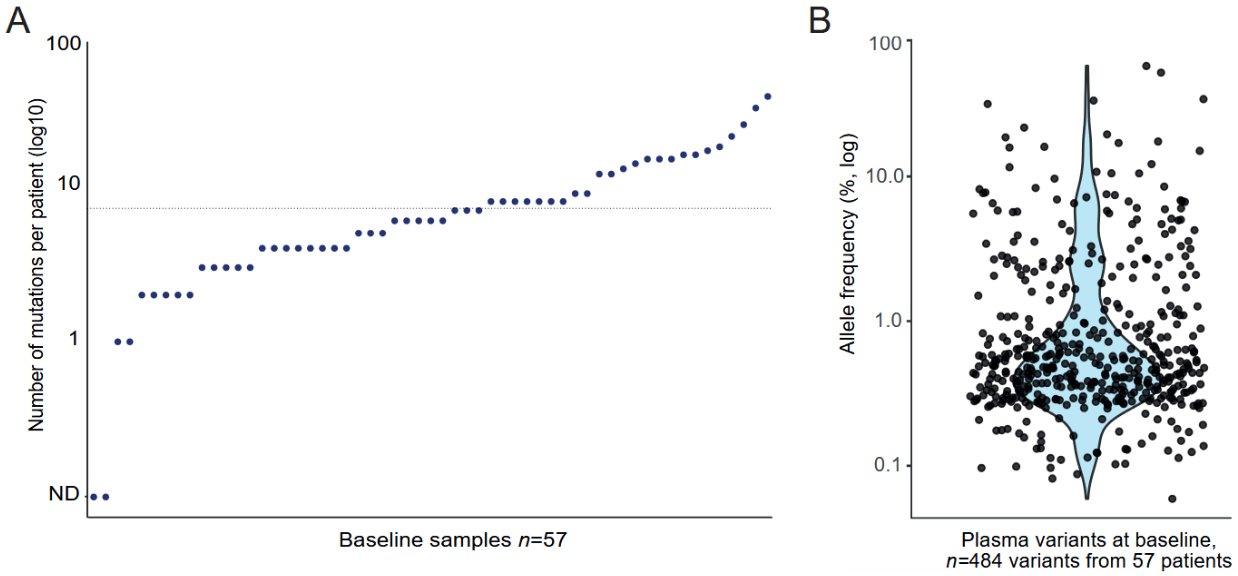


**Supplemental Figure 4. Variant detection in baseline plasma ctDNA. (A)** Number of mutations detected per patient in baseline plasma samples. Each dot represents the number of mutations detected in one baseline plasma sample, ordered from lowest to highest (left to right). **(B)** Violin plot showing the distribution of AFs for all plasma variants identified in baseline samples.

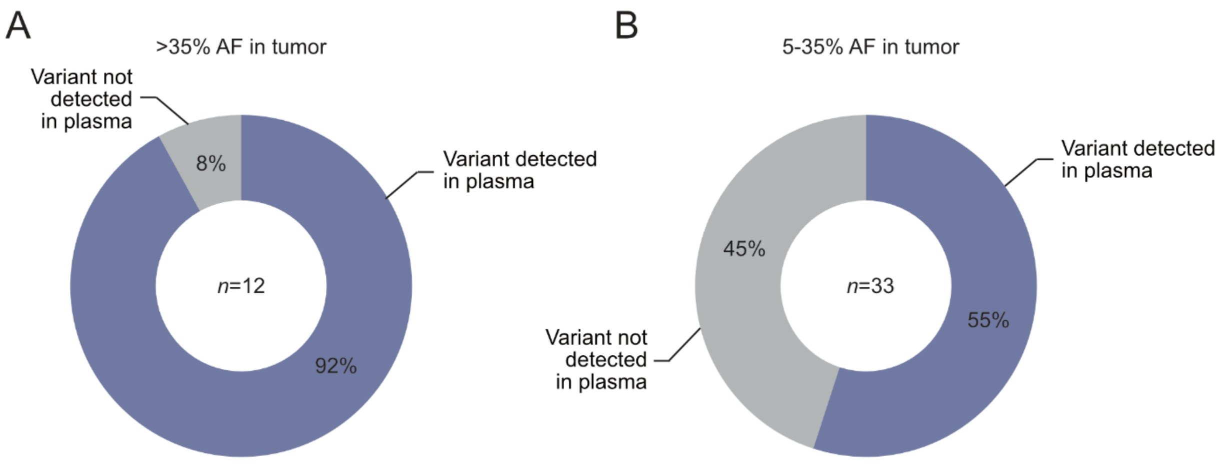


**Supplemental Figure 5. Variant detection of actionable mutations in baseline plasma samples based on allelic representation in tumor tissue. (A,B)** Circle plots showing the proportion of samples with variants detected in plasma, stratified by variant AF in tumor tissue: **(A)** ≥35% and **(B)** 5-35%.

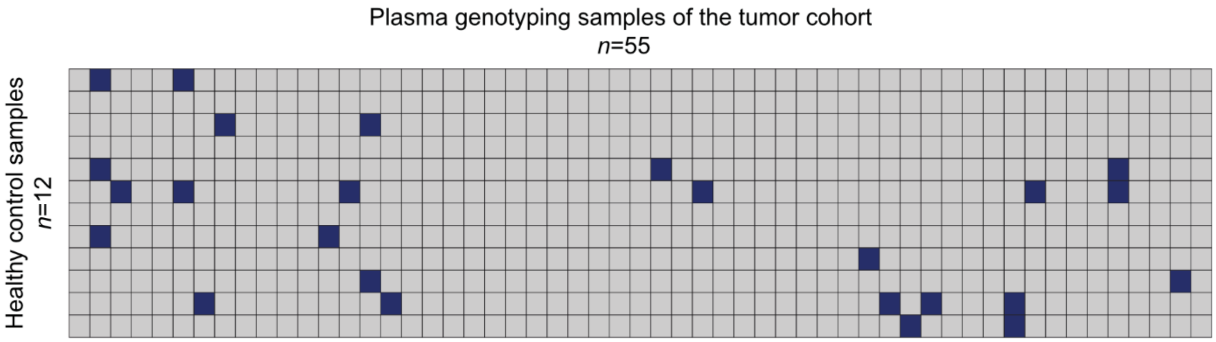


**Supplemental Figure 6. Specificity of the ctDNA monitoring approach.** Heatmap showing the results of the Monte Carlo approach, monitoring baseline plasma ctDNA variants in the withheld cohort of healthy control plasma samples (*n*=12). Grey: true negative, blue: false positive results.

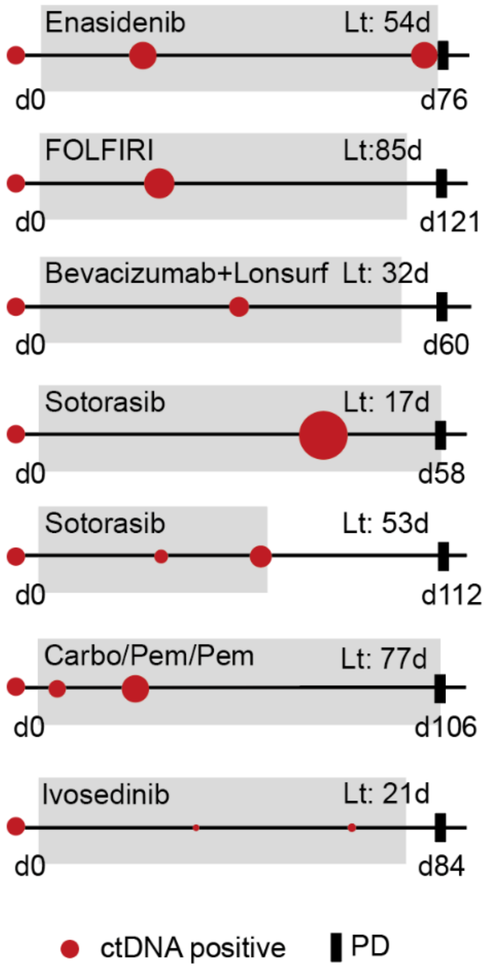


**Supplemental Figure 7. Prediction of disease progression by ctDNA monitoring.** ctDNA dynamics during treatment (grey rectangle) as a function of time. Red dots show positive ctDNA detection. Diameter of the red dots is proportional to relative ctDNA increase/decrease in comparison to the baseline sample (left red dot). Progression time point (PD) is depicted as black rectangle. Lt, lead time; d, day; FOLFIRI, Folinic acid, Fluorouracil, Irinotecan; Carbo/Pem/Pem, combination of Carboplatin, Pemetrexed, Pembrolizumab.

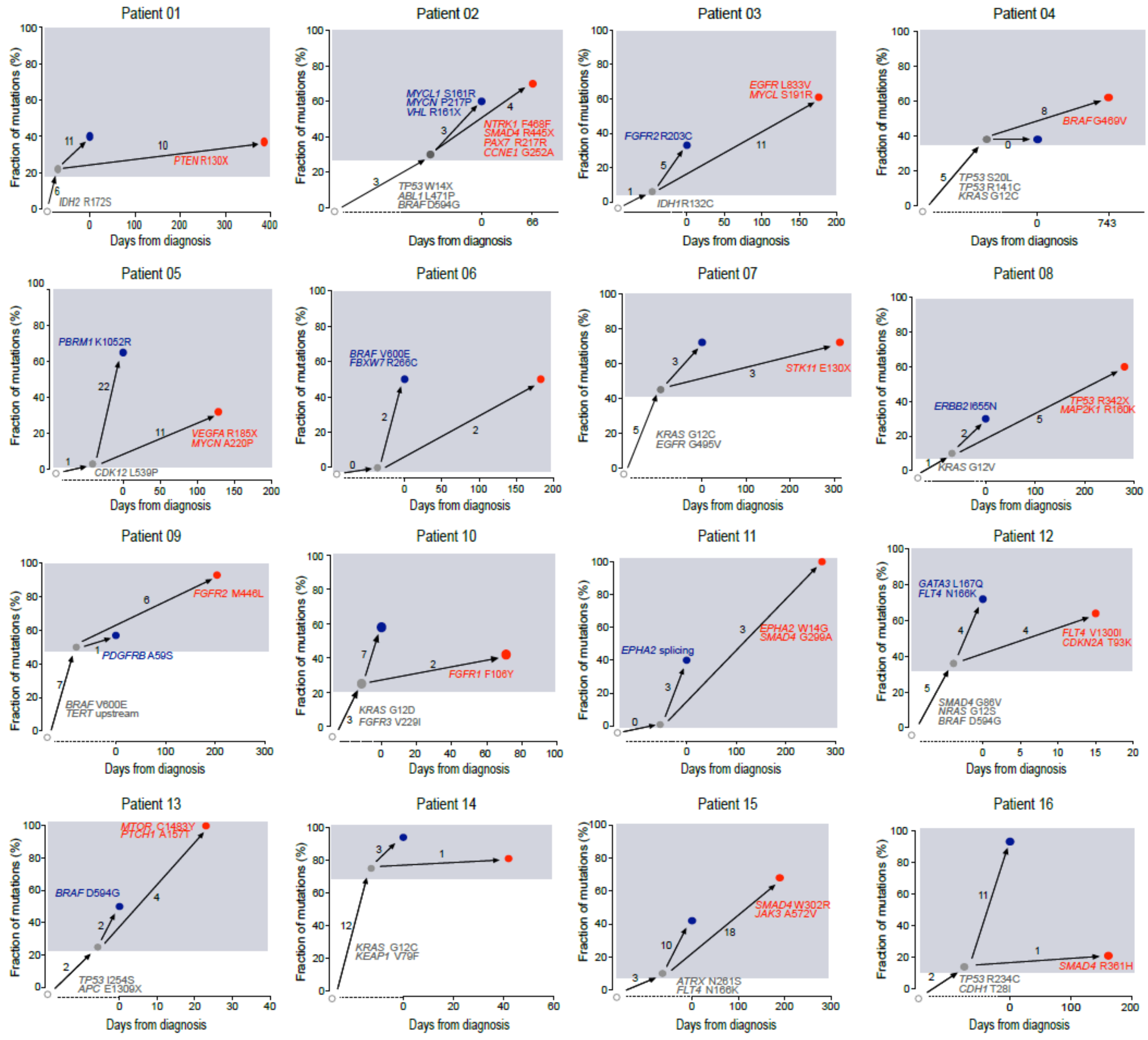


**Supplemental Figure 8. Clonal heterogeneity over time in individual patients identified by ctDNA profiling.** The patterns of clonal evolution in all analyzed 16 cases are shown. The initial branch represents truncal mutations (grey) observed in both plasma samples at baseline and progression. Lost mutations are depicted in blue, emerging mutations in red. The x-axis represents treatment duration, with '0' representing the baseline time point. The y-axis represents the fraction of mutations.

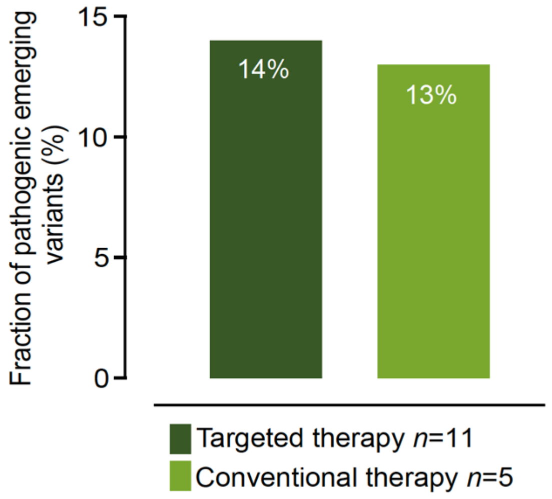


**Supplemental Figure 9. Pathogenic emerging variant across treatment modalities.** Bar graph showing the fraction of pathogenic mutations on all emerging variants per treatment modality.
